# Navigating the Neurological Aftermath of COVID-19: An In-Depth Exploration

**DOI:** 10.1101/2023.09.10.23295343

**Authors:** Maliha Butt, Shavy Nagpal, Ellex Phillips, Shazia Q. Shah, Zeryab Dogar, Hanyou Loh, Sakshi Mishra, Rupalakshmi Vijayan, Rishan Jeyakumar, Sarabjot Singh Makkar, Samia Jahan, Gabriela Marie Díaz, Hudson Franca, Schaza Javed, Marie-Pierre Belizaire, Manoj Reddy Somagutta

**Affiliations:** Division of Clinical & Translational Research, Larkin Health System, South Miami, FL, USA; St Elizabeth Health Centre-Boardman, Ohio

**Keywords:** COVID-19, SARS-Cov-2, Neurological Sequelae, Neurological Manifestations, Acute Neurological Complications, Chronic Neurological Complications

## Abstract

**Background:** The COVID-19 pandemic caused by SARS-CoV-2 has affected millions of people and can result in both immediate and prolonged neurological effects, including severe complications. While numerous studies have explored the occurrence and consequences of neurological issues in COVID-19, they have often involved limited sample sizes.

**Purpose:** This paper aims to determine the overall occurrence of neurological complications in COVID-19, examine their links with patient demographics, and assess their impact on patient outcomes. Additionally, it seeks to provide an overview of the current understanding of the underlying mechanisms.

**Methodology:** Two systematic reviews were conducted to investigate acute and chronic neurological complications associated with COVID-19. A comprehensive search of medical databases was performed, and relevant studies were evaluated following PRISMA guidelines. Meta-analysis was carried out using the Mantel-Haenszel method, with subgroup analysis and meta-regression used to assess heterogeneity.

**Results:** The analysis of acute complications included 20,011 patients with an average age of 58.1 years and a slight male predominance (55.2%). Common neurological symptoms included loss of taste and smell, headaches, acute encephalopathy, and stroke. For the analysis of long-term complications, 2,094 patients were included. Survivors of COVID-19 experienced ongoing neurological issues ranging from sensory impairments to fatigue, headaches, strokes, and even cognitive and psychiatric problems.

**Conclusion:** By examining various neurological symptoms, this study found a significant association between these manifestations and worse overall outcomes, especially in patients over 60 years old. Identifying high-risk individuals and maintaining a high level of suspicion are crucial for enhancing our understanding of the underlying mechanisms, validating biomarkers, and improving the management of these neurological issues.

## INTRODUCTION

COVID-19 has raised numerous questions about its impact on pre-existing health conditions, multi-organ complications, and long-term effects on various bodily systems. Notably, the nervous system has emerged as a particularly concerning area of concern. The spectrum of neurological effects associated with COVID-19 spans from mild issues like changes in taste and smell to severe conditions such as acute encephalopathy, cerebral hemorrhage, and acute myelitis. Patients who experience severe neurological complications appear to face increased risks in terms of both mortality and the severity of their illness. Furthermore, a noteworthy proportion of COVID-19 survivors seem to develop neuropsychiatric disorders, neurodegenerative conditions, and neuropathies in the weeks to months following their acute illness. The involvement of the nervous system during a SARS-Cov-2 infection has thus been linked to substantial morbidity and mortality.

Although several observational studies conducted in clinical settings have examined the neurological effects of COVID-19 and their impact on patient outcomes, many of these studies have been limited by their small sample sizes, making it challenging to generalize their findings to the broader population. In this systematic review and meta-analysis, we aim to aggregate observational studies with similar objectives and structures to establish the incidence of neurological complications associated with COVID-19. Additionally, we seek to explore the associations between these complications and clinical characteristics while also assessing their influence on patient outcomes. This review provides an overview of common neurological manifestations of COVID-19 and endeavors to synthesize our current understanding of the pathophysiology and epidemiology of neurological complications in the context of this viral infection.

## METHODS

We conducted a systematic review following the Preferred Reporting Items for Systematic Reviews and Meta-analyses (PRISMA) guidelines. Our study protocol was registered on PROSPERO with the ID CRD42020209734 on September 22, 2020. To gather relevant literature, we conducted an extensive search on databases including PubMed, Scopus, Google Scholar, and ScienceDirect.

For the analysis of acute neurological complications associated with COVID-19, our search strategy involved various combinations of keywords such as “COVID-19,” “neurology,” “nervous system,” “neurological manifestation,” “acute complications,” and “neurological sequelae.” Additionally, we reviewed the reference lists of existing review articles and conducted manual searches to identify any additional pertinent studies.

Our inclusion criteria encompassed the following: (1) Original studies, such as cohort studies, case-control studies, and cross-sectional studies, (2) confirmation of COVID-19 through laboratory diagnosis in the study population, (3) studies reporting data on neurological manifestations, symptoms, or complications in COVID-19 patients, (4) inclusion of adult patients (age >18 years), and (5) articles published in the English language. We excluded studies falling into the following categories: case reports, case series, non-original articles like review articles (including meta-analyses), letters, or comments.

For our analysis of long-term neurological complications of COVID-19, we manually searched the medical literature using various search terms, such as “chronic neurological complications,” “long-term complications,” “COVID-19,” and “COVID-19 long haulers.” We selected articles based on the following criteria: (1) Observational studies, including cohort studies, case-control studies, and cross-sectional studies, (2) inclusion of patients with laboratory-confirmed COVID-19, (3) study participants limited to adults (age >17 years), and (4) a follow-up period of at least one month after the initial infection. We excluded articles not written in English, as well as case reports, case series, non-original articles like review articles (including meta-analyses), letters, or comments.

### Risk Assessment

We evaluated study quality using the National Institutes of Health (NIH) and National Heart, Lung, and Blood Institute’s (NHLBI) Quality Assessment Tool, along with the Cochrane Risk of Bias assessment tool. Two independent investigators assessed each study’s quality, looking for flaws in design or implementation. Key aspects assessed included clear objectives, well-defined study populations, adequate participation rates, similar patient recruitment, sample size justification, appropriate exposure measurement, sufficient time frames, clear outcome definitions, blinding, minimal loss to follow-up, and adjustment for confounders. Ratings were recorded for each aspect, and an overall study rating was assigned. In cases of disagreement, reviewers discussed the article to reach a consensus, with a third author reconciling differences if necessary.

### Risk of Bias Assessment

We jointly assessed bias using Cochrane’s Risk of Bias tool for Single Studies, considering multiple potential bias domains, such as random sequence generation, allocation concealment, blinding of participants and outcomes, incomplete outcome data, and selective reporting. A low or high-risk rating was assigned to each domain, allowing an overall risk level to be determined based on these judgements. Studies with high bias risks were considered for exclusion.

### Data Extraction

Data extraction was conducted using the Google Sheets app. Initially, a pilot extraction sheet was created and later modified for final review. Two separate groups of co-authors independently reviewed included articles for necessary information. Missing data was requested from corresponding authors, and any disagreements between reviewers were resolved through discussion and consensus.

### Data Synthesis

Meta-analysis was performed using Cochrane Review Manager (version 5.4) and the Mantzel-Haenszel statistical method. A random-effects model was applied to account for inter-study variability. Only studies reporting outcomes in both patients with and without neurological manifestations were included. Composite poor outcomes, including severe/critical COVID-19 infection, ICU admission, mechanical ventilation, and mortality, were analyzed. Risk ratios with 95% confidence intervals were reported for outcomes in patients with neurological manifestations compared to those without. Subgroup analysis based on age (average age below or above 60 years) was conducted to explore sources of heterogeneity. Results were displayed in a forest plot with effect sizes, and a funnel plot was considered if the minimum number of studies for a meta-analysis was reached. Additionally, univariate meta-regression was performed to identify confounding factors affecting study outcomes.

## RESULTS

We identified 350 articles from medical literature databases, journals, and reference lists. After removing duplicates, 296 original articles remained. Through sequential screening of abstracts and full-text articles, we selected 32 eligible studies. Eight of these studies had a high risk of bias and were excluded from the final analysis. Ultimately, 26 studies, primarily cohort studies (16), along with some observational studies and one case series, were included in the systematic review. All these studies collected patient data between January and November 2020.

For the analysis of long-term neurological effects of COVID-19, we conducted a thorough manual search based on the mentioned criteria. After reviewing available studies, we incorporated six cohort studies into our analysis. The limited amount of data currently accessible in this area is reflected in the small number of articles included.

**Table 1:** Demographics [1-6].

| **Demographics** | **Acute Neurological Complications** | **Long-Term Neurological Complications** |
| --- | --- | --- |
| **Total Patients Analyzed** | 20,011 | 2,094 |
| **Average Age** | 58.1 years | 55.7 years |
| **Gender Distribution** | Male: 55.2% | Male: 51.0% |
| **Geographic Distribution (%)** | North America: 57.3% | - |
|  | Europe: 30.2% | - |
|  | Asia: 10.3% | - |
|  | Africa: 0.5% | - |
| **Hospitalized Patients (%)** | 97% | - |
| **Severity of COVID-19 (%)** | Severe to Critical: 28.5% | Severe: 62.1% |
|  |  | Mild/Moderate: 30.2% |
|  |  | Critical: 6.7% |
| **Common Comorbidities (%)** | Hypertension: 39.7% | Hypertension: 26.4% |
|  | Diabetes: 28.8% | Diabetes: 11.7% |
|  | Heart Disease: 28.0% | Heart Disease: 7.9% |
|  | - | COPD: 3.6% |
|  | - | Malignancy: 2.4% |
|  | - | Neurological Disorders: 2.2% |
|  | - | CKD: 2.0% |
| **Common Presenting Symptoms (%)** | Fever: 56% | Insomnia: 22.2% |
|  | Cough: 55% | Mood & Anxiety Disorders: 17.5% |
|  | SOB: 46.2% | Anosmia: 14.7% |
|  | Fatigue: 43.6% | Headache: 9.36% |

| Demographics | Acute Neurological Complications | Long-Term Neurological Complications |
| --- | --- | --- |
|  | Gastrointestinal Symptoms: 39% | Ageusia: 7.31% |
|  |  | Dizziness: 5.97% |
|  |  | Numbness/Tingling, Tinnitus/Blurred |
|  |  | Vision (Both at 1.43%) |
| Quality of Life Assessment | EQ-5D-5L, SF-36, PHQ-9, GAD-7, SRGQ | EQ-5D-5L, SF-36, PHQ-9, GAD-7, SRGQ |
| Cognitive Function Assessment | NIH Toolbox | NIH Toolbox |
|  | Attention: Median T-score 41.5 | - |
|  | Working Memory: Median T-score 43 | - |

### Quality Assessment

Acute Neurological Complications:

- 24 out of 32 studies were of good quality and low risk of bias.
- One study with low risk of bias was excluded due to a small eligible population.
- 7 studies had a high risk of bias and were excluded.

Long-Term Neurological Complications:

- 6 out of 7 long-term studies were of good quality and lower risk of bias.
- One study of poor quality with potential biases in data presentation was excluded.

**Table 2:** Acute Neurological Complications.

| Complication/Manifestation | Incidence Rate (%) | Mean Delay (Days) |
| --- | --- | --- |
| Ageusia | 27.4% | 11.3 (Range: 2 - 28) |
| Anosmia | 21.00% |  |
| Headache | 17.66% |  |
| Delirium/Acute Encephalopathy | 10.99% |  |
| Stroke | 3.75% |  |
| Peripheral Nervous System (PNS) Disease | 3.16% |  |
| Coma | 1.69% |  |
| Seizures | 1.44% |  |

**Table 3:** Association Between Neurological Complications and Poor Composite Outcome.

| Analysis | Risk Ratio (95% CI) | Heterogeneity (I<sup>2</sup>) | Subgroup Analysis |
| --- | --- | --- | --- |
| Neurological Complications vs. No Neurological Complications | 2.06 (1.57 - 2.70) | 94% | Based on Average Age: Older and Younger than 60 Years |
|  |  |  | - Older than 60 Years: Z value: 6.45, P < 0.00001 |
|  |  |  | - Younger than 60 Years: Z value: 1.21, P < 0.23 |
|  |  |  | - Age Subgroup Analysis Did Not Explain Heterogeneity |

**Table 4:** Mortality in COVID-19 Patients with Neurological Manifestations.

|  | Risk Ratio (RR) | 95% CI | P-Value |
| --- | --- | --- | --- |
| Overall | 1.14 | 0.80 - 1.63 | 0.47 |
| Subgroup Analysis |  |  |  |
| Age > 60 Years | 1.75 | 1.14 - 2.70 | 0.01 |
| Univariate Analysis |  |  |  |
| Age | N/A | N/A | 0.754 |
| Hypertension | N/A | N/A | 0.449 |
| Diabetes Mellitus | N/A | N/A | 0.228 |
| Hyperlipidemia | N/A | N/A | 0.185 |
| Pre-existing Neuro | N/A | N/A | 0.501 |
| Cancer | N/A | N/A | 0.294 |
| COPD | N/A | N/A | 0.686 |
| CKD | N/A | N/A | 0.391 |
| Heart Disease | N/A | N/A | 0.463 |
| Heterogeneity (I<sup>2</sup>) | 90% |  |  |

## DISCUSSION

Neurological complications in COVID-19 are diverse, ranging from mild symptoms like headache and anosmia to severe conditions including encephalitis and stroke. These complications can persist long after the acute infection. The overall incidence is around 25.2%. Severe neurological issues are less common than cardiovascular problems but can have significant morbidity and mortality.

Gustatory dysfunction (GD) and anosmia are common symptoms, often occurring together. These symptoms can vary in prevalence across regions and may be underreported in severe cases.

Delirium and acute encephalopathy are significant concerns, especially in older patients, and can be early signs of COVID-19. They are associated with adverse outcomes and increased mortality. Management includes identifying high-risk patients and addressing underlying causes, with steroids showing promise in treatment. [7-56]

Stroke in COVID-19 is a rare but serious complication, often occurring after the first week of infection. It’s associated with endothelial damage and inflammation due to the virus, leading to large artery strokes. The incidence is around 3.86%, with a higher risk in older, male, and comorbid individuals.

Seizures can occur in COVID-19, especially in serious cases, and may be due to various causes, including fever, hypoxia, and underlying conditions. The incidence is approximately 1.44%. Coma in COVID-19 is associated with poor outcomes, especially in patients over 60. It can result from various factors, including sedation in the ICU and underlying encephalopathy. The incidence is around 1.69%. [57-88]

Headache is a common neurological symptom in COVID-19, possibly caused by direct virus interaction with the nervous system or inflammation-related factors. It’s among the top symptoms after fever, cough, myalgia, and dyspnea.

The incidence of headache in COVID-19 varies, with rates from 5% to 37% in different studies. Anosmia and ageusia are often associated with headaches.

COVID-19-related headaches can have various characteristics, including migraine-like features and unusual aspects like intractable headaches. Some patients experience persistent or late-onset headaches after the acute phase of the illness.

Treatment typically starts with acetaminophen and may progress to NSAIDs if necessary. Refractory headaches may require more specialized evaluation. In some cases, headaches can persist for several months as a long-term symptom of COVID-19. [89-105].

COVID-19 is associated with various peripheral nervous system (PNS) complications, including neuralgias, Guillain-Barre Syndrome (GBS), cranial nerve disorders, neuro-ophthalmological issues, sensorineural hearing loss, and autonomic dysfunction.

The incidence of PNS disorders in COVID-19 varies widely, with reports ranging from 9% to 60%. In our analysis, PNS disorders occurred in 3.56% of COVID-19 patients, with peripheral neuropathies and cranial nerve disorders being the most common.

PNS complications may result from direct viral invasion or systemic inflammation. COVID-19-related GBS is more common in older patients and those with severe disease. It can precede other COVID-19 symptoms and often has a poor prognosis.

Dysautonomia, characterized by blood pressure instability, orthostatic hypotension, and bladder/bowel dysfunction, is common in critically ill COVID-19 patients and may be related to nerve pathway disruption.

New-onset neuromuscular diseases like Myasthenia Gravis have been reported in COVID-19 patients, possibly due to molecular mimicry.

Treatment for PNS complications varies based on the diagnosis but often includes steroids, plasma exchange, and immunoglobulins for GBS and related disorders. Opioids and anticonvulsants should be avoided for neuralgic pain in COVID-19 patients. [106-119]

### Fatigue

Fatigue and myalgia are common post-SARS-CoV-2 infection symptoms, with varying prevalence rates in studies. Factors such as gender, disease severity, age, BMI, and pre-existing depression may influence the likelihood of experiencing long-term fatigue.

The mechanisms behind acute and chronic fatigue involve immune and nervous system components, including neuroinflammation and endothelial cell damage. There is no definitive causal mechanism for chronic fatigue, and further research is needed to better understand its etiology, clinical course, and treatment. [120-154].

## LIMITATIONS OF THE STUDY

This systematic review analyzes the impact of individual neurological complications on COVID-19 mortality. Caution is needed due to the limited number of studies, potential bias, and lack of control groups. Only stroke and acute encephalopathy were included in the meta-analysis. Severe COVID-19 may contribute to severe neurological disorders, but causal relationships need further investigation. Future studies should address confounding factors and clarify COVID-19’s relationship with neurological complications.

## CONCLUSION

Acute neurological disorders in COVID-19 are common, associated with poor outcomes and higher mortality, especially in those over 60 years. Survivors can experience chronic complications, including sensory issues, fatigue, stroke, cognitive, and psychiatric problems. Ongoing research, like the BRAINSTORM study, is crucial for better understanding and targeted management of these conditions. Healthcare workers should maintain a high index of suspicion for early detection.

## Data Availability

All data produced in the present work are contained in the manuscript

